# Wearable Myoelectric Interface for Neurorehabilitation (MINT) to Recover Arm Function: a Randomized Controlled Trial

**DOI:** 10.1101/2025.06.24.25330240

**Authors:** Abed Khorasani, Cynthia M. Gorski, Na-Teng Hung, Joel Hulsizer, Vivek Paul, Goran Tomic, Prashanth R. Prakash, Sangsoo Park, Gang Seo, Ethan J. Houskamp, Jessica Lanis, Megan Hunzeker, Erin King, Anya Chappel, Alix Jampol, Pooja Patel, Catherine Gallagher, Rachel Galant, Grace Rucker, Jungwha Lee, Richard Harvey, Jinsook Roh, Marc W. Slutzky

**Author notes:** Corresponding Author: Marc W. Slutzky, Department of Neurology, 320 East Superior Ave., Searle 11-473, Chicago, Illinois, 60611. Co-Corresponding Author: Jinsook Roh, Department of Biomedical Engineering, 3517 Cullen Blvd., Rm 2027, Houston, Texas, 77204-5060, Total words: 4998.

## Abstract

**Background:** Abnormal muscle co-activation contributes to arm impairment after stroke. This single-blind, randomized, sham-controlled trial evaluated the feasibility and efficacy of home-based, personalized myoelectric interface for neurorehabilitation (MINT) conditioning to reduce abnormal co-activation and enhance arm function and determine the optimal number of abnormally co-activating muscles to target during training.

**Methods:** Moderately to severely impaired chronic stroke survivors were randomized to one of three MINT groups (who played customized games requiring independent activation of 2 or 3 abnormally co-activating muscles) or a sham control group (played using one muscle). All groups trained 90 minutes/day, 5 days/week at home and 1 day/week in lab, for 6 weeks, and changed trained muscle sets every 2-3 weeks. The primary outcome was the Wolf Motor Function Test (WMFT) at 6 weeks.

**Results:** Fifty-nine participants completed the training. Participants performed 315 ± 85 (mean ± SD) repetitions daily. At week 6, participants in all MINT groups combined improved by 4 s on WMFT (p=0.0008), exceeding the minimal clinically important difference (1.5 s). Participants who trained 3 muscles simultaneously improved by 6.8 s (p=0.001), while the 2-muscle and sham groups did not change significantly. In per-protocol analysis, the 3-muscle group, but not 2-muscle groups, improved significantly more than sham (p=0.046), though not in intention-to-treat analysis. All MINT groups continued improving at 4 weeks post-training. Importantly, severely impaired participants in combined MINT groups improved more than those in sham (p=0.02). Importantly, combined MINT groups also improved their reaching range of motion significantly more than sham. Co-activation decreased by 76% in MINT groups during training. Notably, reduction in co-activation during reaching correlated significantly with improved arm function and range of motion. Other secondary outcomes did not show clinically important improvement. Stroke involving the posterior limb of the internal capsule negatively predicted response to MINT.

**Conclusions:** Home-based MINT conditioning, especially the 3-muscle variant, is feasible, reduces co-activation, and improves arm movement and function.

**Clinical Trial Registration:** ClinicalTrials.gov (NCT03401762)

## Introduction

Stroke is a leading cause of permanent disability worldwide, with about 60% of survivors experiencing impairment in upper limb function six months after stroke^1,2^. Although rehabilitation therapies can improve function even after the initial recovery period^3–5^, only 30% of stroke survivors in the U.S. receive outpatient therapy due to resource constraints^6^. Further, higher doses of therapy are important in therapeutic improvement^7^. These facts highlight the need for an effective, affordable, and widely accessible stroke treatment.

Arm impairment following a stroke is not just due to weakness and spasticity but also due to impaired joint coordination from abnormal muscle co-activation, also called abnormal muscle synergies^8,9^, which is not directly addressed by existing therapies. Our initial studies on chronic stroke survivors showed that in-lab training with a myoelectric computer interface that provided visual feedback about abnormal arm muscle co-activation reduced co-activation and associated arm impairment^10–12^. Further, a higher dose of training (90 min/day) while moving led to better outcomes than isometric training^11^.

In the current study, we used a wearable version of the myoelectric interface, called myoelectric interface for neurorehabilitation (MINT), in a home-based, randomized, sham-controlled trial. MINT delivers personalized gamified training tailored to each patient’s co-activation patterns. It could ultimately provide a low-cost addition to conventional therapy and an option for individuals with limited access to outpatient rehabilitation.

We sought to further optimize the MINT paradigm for the upper arm in terms of the number and ways that muscles were trained (2 vs. 3 muscles simultaneously, with or without visual cues), and to validate that it was doing more than overcoming non-use. Thus, the goals of this clinical trial were to determine 1) if MINT at home can improve arm function in chronic stroke survivors, 2) which variant of the MINT paradigm was effective (similar to a dose-finding design^13^), and 3) whether reducing abnormal co-activation leads to improved arm motor function by means other than overcoming non-use of the arm.

## Methods

### Participants

This study follows the CONSORT guidelines (Consolidated Standards of Reporting Trial Statement). The study (NCT03401762), approved by the Institutional Review Board of, and conducted at, Northwestern University, aimed to assess feasibility and the effect of MINT conditioning on arm function. Written informed consent was obtained from all participants prior to eligibility assessment. Recruitment was conducted through multiple channels, including the clinical neuroscience research registry, day rehabilitation centers, hospital websites, and physician referrals. Participants were required to be over 18 years old, with moderately severe to severe upper limb impairment, indicated by Fugl-Meyer Assessment of Upper Extremity (FMA-UE) scores from 7 to 30, following a unilateral ischemic or hemorrhagic stroke occurring at least 6 months before enrollment. Study candidates were excluded for significant visual deficits impairing visibility of a laptop screen (mainly visual field deficits), language comprehension deficits (assessed by 3-step commands and ability to understand the study), bilateral strokes, recent botulinum toxin treatment in the affected arm within 3 months of screening, new physiotherapy initiation within 3 months of screening, arm contractures, or participation in other upper limb-related research studies within 3 months of enrollment. Participants were randomized into one of 3 experimental groups (see next section) or a sham group, with stratification based on age and baseline impairment level. A randomization schedule was generated by a biostatistician using R software, and group assignments were carried out by a team member who had no interaction with, nor involvement in the evaluation of the participants.

### Identifying Abnormal Co-activation

Two weeks before training, participants were screened in the laboratory for abnormal co-activation using a seated reaching task (Fig. S1 A). This task involved reaching as far as possible three times each toward six targets in front and six targets to the side, positioned at waist and shoulder height, totaling 36 reaches (Fig. S1 A). During reaching, surface electromyographic (EMG) was recorded from 12 arm and shoulder muscles of the affected arm: brachioradialis (BRD), biceps brachii (BI), triceps long (TRIlong), lateral triceps (TRIlat), anterior deltoid (AD), middle deltoid (MD), posterior deltoid (PD), pectoralis major (Pec), trapezius (Trap), infraspinatus (Inf), latissimus dorsi (Lat), and pronator quadratus (PQ) using active sensors (Trigno, Delsys, Inc.). The muscles exhibiting the highest abnormal co-activation, determined by pairwise correlation coefficients, were identified to customize the training muscle sets for each participant (Fig. 1S B). Co-activation between the following muscle sets were considered abnormal^15^: anterior deltoid/biceps, anterior deltoid/posterior deltoid, anterior deltoid/brachioradialis, biceps/triceps, brachioradialis/triceps, pectoralis major/biceps, pec major/brachioradialis, pronator/biceps, anterior deltoid/trapezius. As mentioned above, we sought both to compare to a control intervention and to optimize the design of MINT conditioning. Participants were randomized into one of four groups (Fig. 1B, Fig. S1): (1) 2D group, who trained with MINT using 2 muscles at a time to control a cursor (see next section for details); (2) 2D Reach group, who also used 2 muscles simultaneously to control the cursor while being cued with pictures instructing them to reach as far as possible in the direction of the target muscle; (3) 3D group, who used 3 muscles simultaneously to control the cursor; and (4) sham group, who used one muscle to control a cursor in 1D. We hypothesized that the 2D Reach condition was a context more similar to normal arm use and therefore might induce better transfer of learning to arm function in daily use^16,17^. Similarly, decoupling 3 muscles at a time might also be a context more similar to daily use than just 2 at a time, and therefore might transfer better to daily use^16,17^. The sham group controlled for the possibility of MINT improving function due to overcoming non-use of the impaired arm. Muscle sets were changed every 2 weeks in 2D/2D Reach groups and 3 weeks in the 3D group (Fig. S1). This choice was intended to approximately balance the total number of muscles trained over 6 weeks in experimental groups. The sham group also changed muscles every 2 weeks. Participants were not aware of what the difference in training was between their group and other possible groups. The list of trained muscles for all participants is reported in Table S1.

**Figure 1.**
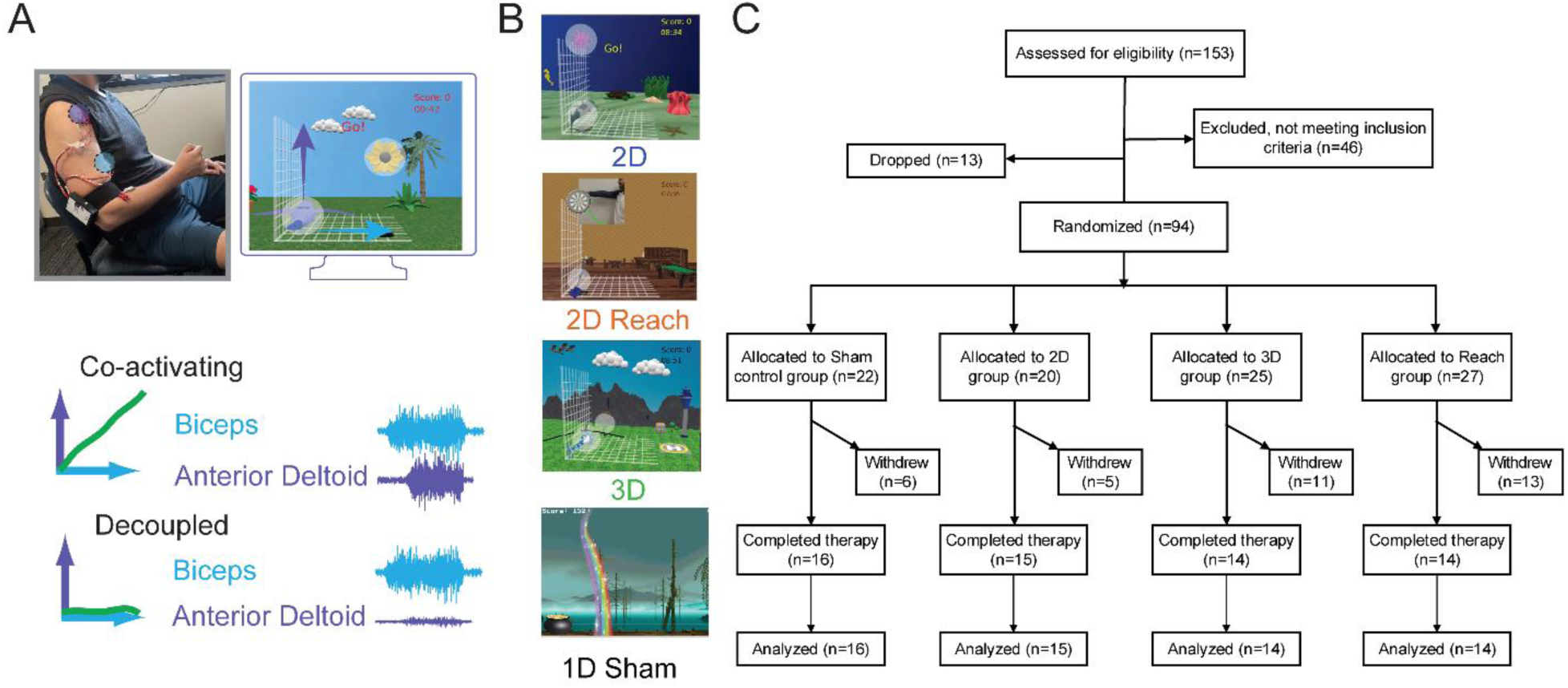
MINT paradigm and CONSORT diagram. (A) *Top*, an example stroke participant used the wearable MINT device to transmit EMGs of biceps (blue) and anterior deltoid (purple) to the laptop. *Bottom*, MINT software mapped EMGs of these muscles to orthogonal components of cursor movement. In the example, biceps was mapped to the right (blue arrow) and anterior deltoid was mapped up (purple arrow). When biceps and anterior deltoid were co-activated, the cursor moved along a diagonal between the two directions (green). The participant was then conditioned using MINT to decouple the two muscles; in this example, to activate biceps independently of anterior deltoid. (B) Examples of MINT game screens for the four groups. In the 2D group, participants used two muscles mapped to the x and y directions. Similarly, in the 2D Reach group, participants were trained with two muscles, but they were given additional visual prompts to encourage more effort to reach in the current muscle’s pulling direction. In the 3D group, participants used three muscles simultaneously, with each muscle separately mapped to the x, y, and z directions of cursor movement. Finally, in the 1D sham group, participants used only one muscle mapped to the x direction. (C) CONSORT enrollment flow chart.

### MINT Conditioning

Participants were asked to use the MINT system for 90 minutes per day, five days a week at home, and one day a week in the lab, over six weeks. Each training session was divided into nine “runs” of 10 minutes each. On average, each run consisted of about 30 trials (repetitions), which were randomly ordered and balanced over target muscles. MINT included a custom-built, wireless surface EMG acquisition system (Myomo, Inc.) that amplified, digitized, and computed the envelope of the EMGs, and transmitted the envelopes at 50 Hz to a laptop via Bluetooth.

In the MINT game, participants manipulated a cursor within a customized three-dimensional environment created using Blender and Python (Fig. 1A). The EMG envelopes were mapped to orthogonal components of cursor position in either 2D (for 2D and 2D Reach groups) or 3D (for 3D group). The vector sum of these components determined the cursor position at each 50 Hz sample. To reach targets on the cardinal axes, participants in the experimental groups had to activate the targeted muscle in isolation from other muscles. Participants initiated cursor movement into the “home” target at the bottom left of the screen by relaxing the controlling muscles (Fig. 1A). Upon successful arrival at the home target, an outer target appeared near one of the neighboring corners of the screen, requiring participants to activate the controlling muscle to move the cursor into the target and maintain its position for 0.5 seconds. Positive audiovisual feedback was provided for successful trials, while failure to acquire the target within 7 seconds resulted in an unsuccessful trial. Detailed descriptions of the MINT game design and difficulty levels were published previously^18^ and more details can be found in Supplementary Methods.

### Outcome Measures

Behavioral metrics during training, including weighted time to target (the time to acquire a target for each trial, a measure of game performance) and co-activation between trained muscles, were computed for each 10-min run. To account for the level of difficulty during MINT training, the time to target obtained from both successful and failed trials was normalized by the difficulty level (by dividing by the level) for each run. Co-activation between trained muscles from both successful and failed trials was computed in a 1-s temporal window before target capture. Game performance was computed for the two weeks trained on each muscle set separately for the 2D, 2D Reach, and sham groups; co-activation was similarly computed for each muscle set in both 2D groups. Since the sham group only used one muscle at a time and only one was recorded, the co-activation metric could not be computed. For the 3D group (those who trained for 3 weeks on each muscle set) only data from the first two weeks of training on each set were reported to facilitate aggregation of metrics with other groups.

Evaluations were conducted by an occupational therapist blinded to group assignment at −2, 0, 2, 6, and 10 weeks relative to the initiation of training (Fig. S1). The primary clinical outcome was the change in the timed portion of the Wolf Motor Function Test (WMFT) at the end of training (Week 6) relative to the baseline (the mean of Week −2 and Week 0). We chose WMFT because, as a continuous (timed) measure, it is more sensitive to change than other discrete metrics such as the Fugl-Meyer Assessment of the Upper Extremity (FMA-UE). It also has more items relevant to upper arm function that are independent of hand function than some other clinical scores, such as the Action Research Arm Test. We considered these properties important because this trial was a) enrolling mostly severely impaired participants who had little or no hand function and b) only training muscles related to elbow and shoulder function. Secondary clinical outcomes included the WMFT at Week 10 (4-week follow-up), shoulder and elbow items of the WMFT, FMA-UE (motor), Modified Ashworth Scale (MAS), and Motor Activity Log (MAL). Occupational therapists were trained to perform these assessments by experienced trainers in a standardized fashion and tested to ensure at least 95% proficiency before being approved as an assessor. In addition, we computed arm kinematics, as described in Supplementary Methods.

Participants’ engagement and effort were assessed using a modified version of the Intrinsic Motivation Inventory (IMI) at the end of the 6-week training period. The IMI questionnaire included 18 questions, each rated on a 7-point Likert scale from 1 (‘Strongly Disagree’) to 7 (‘Strongly Agree’), with 4 indicating neutrality. The questions were adapted from the original IMI to reflect participants’ experiences with MINT conditioning. They were grouped into four categories: enjoyment, effort, perceived benefit, and satisfaction. Mean scores were calculated for all participants, overall survey responses, and within each category. For negatively worded questions, scores were subtracted from 8 to compute the means.

### Effect of stroke location on MINT recovery

Lesions were identified by team members with clinical neuroimaging experience. Stroke lesion locations were defined as areas of restricted diffusion in MRIs from the acute stroke phase, if available, or from T2 hyperintensities in MRIs from the chronic phase^20,21^. The lesion locations for all participants are reported in Table S2. Lesion locations were grouped as follows: structures of the corticospinal tract, i.e., the primary motor cortex, corona radiata (CR), and posterior limb of the internal capsule (PLIC); basal ganglia (BG: caudate, putamen, globus pallidus). These groupings were chosen based on brain structures commonly involved in controlling movement and affecting stroke recovery^22^. Based on the study by Shelton et al., we hypothesized that participants with lesions in the PLIC, adjacent corona radiata, and basal ganglia would experience less motor recovery following MINT conditioning^22^. We compared changes in the WMFT at week 2, 6 and 10 between groups based on damage to regions of interest (ROIs). These ROIs included: 1) PLIC±: at least the PLIC, which may also include BG, CR, or both, 2) No PLIC: no PLIC involvement, 3) PLIC + BG + CR: specifically including all three areas, and 4) No PLIC/BG/CR: no involvement of any of the three areas.

### Statistical analysis

The sample size was calculated based on a small set of preliminary data from our prior study^11^, in which the effect size estimate was 0.8. We estimated that, for a power of 0.8 and Bonferroni-corrected alpha of 0.05, we would require 14 participants per group to accommodate 15% attrition rate. All participants who completed the 6-week training were included in the intention-to-treat analysis. Those who finished a minimum of 30 hours of training within a span of fewer than 10 weeks were included in the per-protocol analysis. To evaluate the comparisons within and between groups, linear mixed-effects models (LMEMs) were used with estimated marginal means, which also accounted for repeated measures and missing values. The outcome measures of interest were WMFT, FMA-UE, MAS, and MAL. Fixed effects included group (2D, 3D, 2D Reach, and sham) and random effects included repeated time points (baseline, 6 weeks, and 10 weeks). Additionally, two-sample t-tests were performed to evaluate the significance of changes in kinematic outcomes and to assess the influence of injury location following MINT conditioning.

For the kinematic analysis, five subjects were excluded due to technical issues with kinematic recordings. In the stroke location analysis, twelve subjects were excluded due to inability to access to any of their neuroimaging information. No subjects were excluded from the game performance analysis; however, missing training performance data on MINT training days were filled in using the last observation carried forward method. Muscle synergy analysis is described in Supplementary Methods.

## Results

### Participant enrollment

Between January 2018 and March 2024, 153 participants were screened for eligibility (Fig. 1C). Among them, 46 subjects did not meet the inclusion criteria, and an additional 13 subjects withdrew before the start of training at week 0 (Fig. 1C). Consequently, 94 subjects were randomized into four groups, with 25 allocated to the 3D group, 20 to the 2D group, 27 to the 2D Reach group, and 22 to the sham group. During the course of the study, 35 subjects withdrew before week 6 of training. The main reasons for withdrawal were related to insufficient ability or desire to adhere to the protocol and family or personal issues (Table S3). Ultimately, a total of 59 participants (intention-to-treat population), aged 21-87 years, with a mean duration of 6.4 years since stroke, completed the entire 6 weeks of training and were included in the final analysis. Two participants did not return for the follow-up visit at 4 weeks post-training. Table 1 shows participants’ demographics. Ten participants were excluded for the per protocol analysis (6 due to performing less than 30 hours of training, 4 due to taking more than 10 weeks). The Reach group was significantly older than sham (p=0.02, t-test); there were no other significant differences in ages between groups.

**Table 1.** Participant Demographics.

|  | 3D (n = 14) | 2D (n = 15) | 2D Reach (n = 14) | sham (n = 16) |
| --- | --- | --- | --- | --- |
| Age (years) | 60.6 ± 14.5 | 56.7 ± 8.6 | 64.7 ± 14.9 | 52.6 ± 12.6 |
| Gender, n (%) |  |  |  |  |
| Male | 11 (79) | 9 (60) | 11 (79) | 10 (63) |
| Female | 3 (21) | 6 (40) | 3 (21) | 6 (38) |
| Race, n (%) |  |  |  |  |
| White | 6 (43) | 10 (67) | 8 (57) | 8 (50) |
| Black | 7 (50) | 3 (20) | 6 (43) | 6 (38) |
| Asian | 1 (7) | 2 (13) | 0 (0) | 1 (6) |
| Unknown | 0 (0) | 0 (0) | 0 (0) | 1 (6) |
| Handedness, n (%) |  |  |  |  |
| Right | 13 (93) | 11 (73) | 14 (100) | 13 (81) |
| Left | 1 (7) | 4 (27) | 0 (0) | 3 (19) |
| Affected side, n (%) |  |  |  |  |
| Right | 5 (36) | 7 (47) | 8 (57) | 7 (44) |
| Left | 9 (64) | 8 (53) | 6 (43) | 9 (56) |
| Time post-stroke (yrs) | 5.8 ± 4.7 | 5.7 ± 6.3 | 5.1 ± 6.9 | 7.0 ± 11.6 |
| FMA-UE baseline | 14.0 ± 4.3 | 16.7 ± 5.6 | 16.0 ± 4.2 | 17.7 ± 7.1 |
| WMFT baseline | 96.3 ± 11.7 | 90.1 ± 13.6 | 91.7 ± 10.9 | 90.4 ± 19.2 |
| MAS baseline | 0.8 ± 0.3 | 0.8 ± 0.3 | 0.7 ± 0.4 | 0.9 ± 0.3 |
| MAL-Q baseline | 0.5 ± 0.9 | 0.5 ± 0.4 | 0.3 ± 0.4 | 0.6 ± 0.7 |
| MAL-A baseline | 0.5 ± 0.8 | 0.5 ± 0.4 | 0.3 ± 0.4 | 0.6 ± 0.7 |

Abbreviations: FMA-UE, Fugl-Meyer Assessment for Upper Extremity; WMFT, Wolf Motor Function Test; MAS, Modified Ashworth Scale; MAL-Q, Motor Activity Log - Quality of Movement; MAL-A, Motor Activity Log - Amount of Movement. age, time, and baselines display mean ± SD.

### Training adherence and feedback

Participants trained 86 ± 21 min/day of the instructed 90 min/day: 31% of participants completing the trial trained for at least 90 min/day; 91% trained for at least 60 min/day. Participants in the 3D group performed 287 ± 84 repetitions per day, those in the 2D reach group performed 315 ± 92 repetitions per day, those in the 2D group performed 356 ± 85 repetitions per day, and sham performed 300 ± 65 repetitions per day during MINT conditioning.

The modified IMI survey results showed that 73% of participants who completed the study agreed at least slightly that MINT was enjoyable (Fig. S2). Additionally, 96% of participants agreed at least slightly that they used a high amount of effort during MINT. Mild fatigue and skin irritation were the most common adverse events, occurring in 9 participants. Details of adverse events are listed in Table S4. Participants anecdotally reported positive benefits from the MINT game, such as enhanced muscle engagement, reduced arm tension, and increased awareness of their affected arm. While some challenges with device setup and functionality were noted, the overall feedback highlighted the game’s motivational impact and therapeutic value. Comprehensive feedback from participants is detailed in Table S5. We tracked phone interactions with 35 participants. Most participants required limited contact with lab staff during the week; only 6% required support more than three times per week.

### MINT performance and clinical outcomes

Participants learned to perform MINT conditioning accurately relatively quickly, within about 5 days (Fig. S3). During training, there was a 76% decrease in co-activation relative to the reaching task across all muscle pairs trained with MINT in the experimental groups (Fig. S3).

In the intention-to-treat analysis, participants in all experimental groups combined improved by a mean of 4.1 s in the primary outcome, the WMFT, at the end of training (week 6) compared to baseline (*p*=0.0008, Fig. 2A), exceeding the minimum clinically important difference (MCID) of 1.5 s. The 3D group improved by 6.8 s compared to baseline, while the 2D, 2D Reach, and sham groups did not change significantly from baseline (Fig. 2A, Table 2). By week 10, all experimental groups had improved significantly from baseline, but the sham group had not (Fig. 2A, Table 2). In the per-protocol analysis, the 3D group improved significantly more than sham at week 6; other experimental groups did not (Fig. 2B, Table 2). Because participants were only training upper arm muscles, we also analyzed a subset of WMFT items involving movement of the elbow and shoulder only. In the per-protocol analysis of elbow and shoulder items only, the 3D group improved significantly more than sham at week 6 (by 16.2 s), and combined experimental groups also improved significantly more than sham (Fig. 2C, Table 2). In the intention-to-treat analysis of elbow and shoulder items, the 3D group improved significantly more than sham (by 11.9 s) at week 6. The other changes from baseline in the intention-to-treat analysis were not statistically greater than sham for any experimental group after Bonferroni correction (Table 2). The MCID was exceeded at week 6 by 10 of 14 participants (71%) in the 3D group, 7 of 14 participants (50%) in the 2D Reach group, 8 of 15 (53%) in the 2D group, and 7 of 16 participants (44%) in the sham group. Notably, even individuals with severe impairment (initial FMA<25, 95% of our participants) in the combined experimental groups improved by 4.3 s more than those with severe impairment in the sham group at week 6 (*p*=0.02, two-sample t-test). Moreover, effect sizes were moderate to large at week 6—0.7 and 1.2 in combined experimental and 3D, respectively—and increased at week 10–0.9 and 1.2 in combined experimental and 3D, respectively (Table 3).

**Figure 2.**
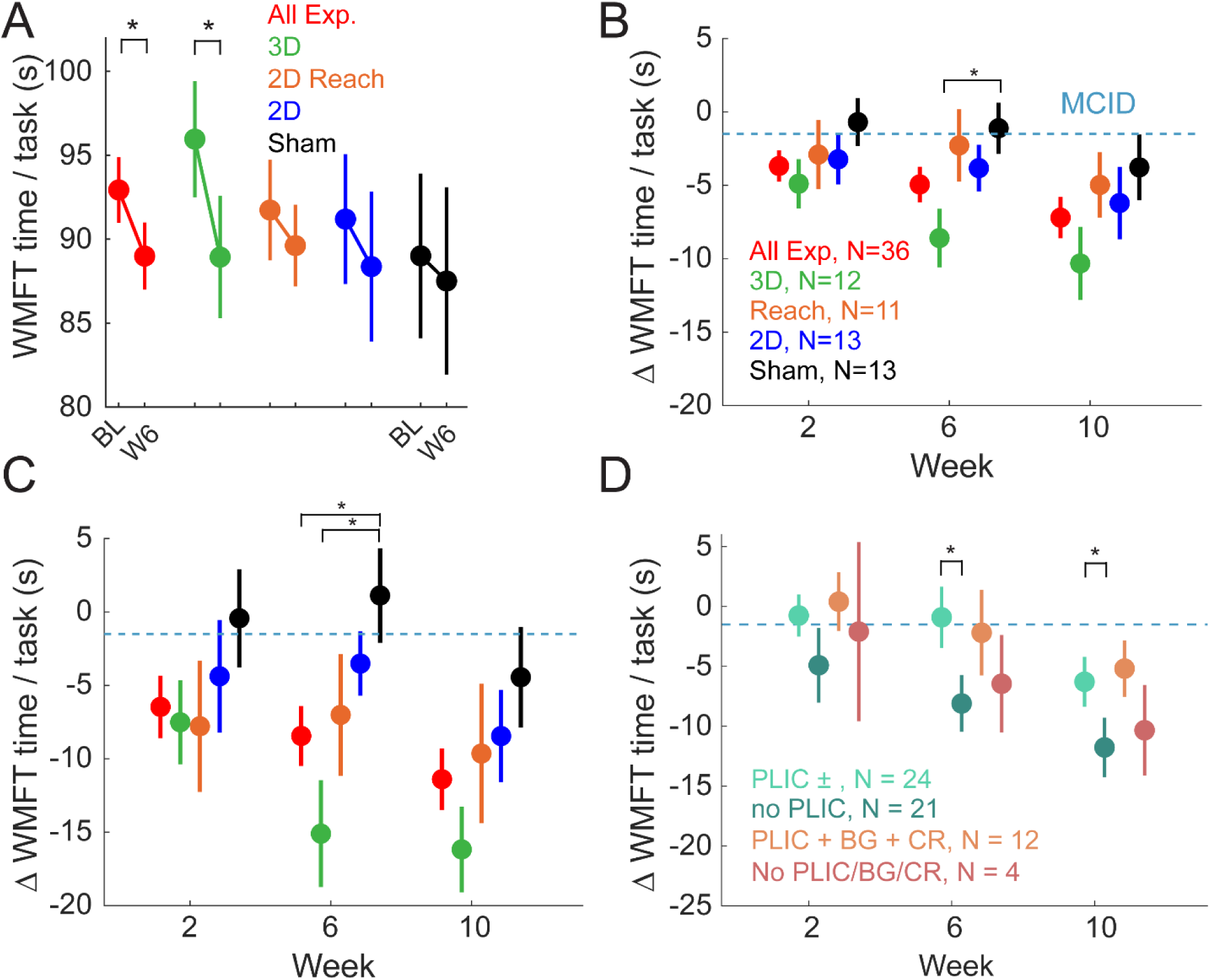
Effect of MINT conditioning on improving motor function. (A) Mean (±SE) WMFT at baseline (BL) and Week 6 (W6) for each group (red, combined experimental groups; green, 3D; blue, 2D; orange, 2D Reach; black, sham). (B) Mean change in WMFT relative to baseline. Dashed line, MCID. (C) Mean change in WMFT relative to baseline for items related only to elbow or shoulder. (D) Mean change in WMFT relative to baseline stratified by injury location for all participants. Abbreviations refer to strokes involving: PLIC, posterior limb of the internal capsule, BG, basal ganglia, CR, corona radiata. Asterisks (*) indicate p < 0.05.

**Table 2.**
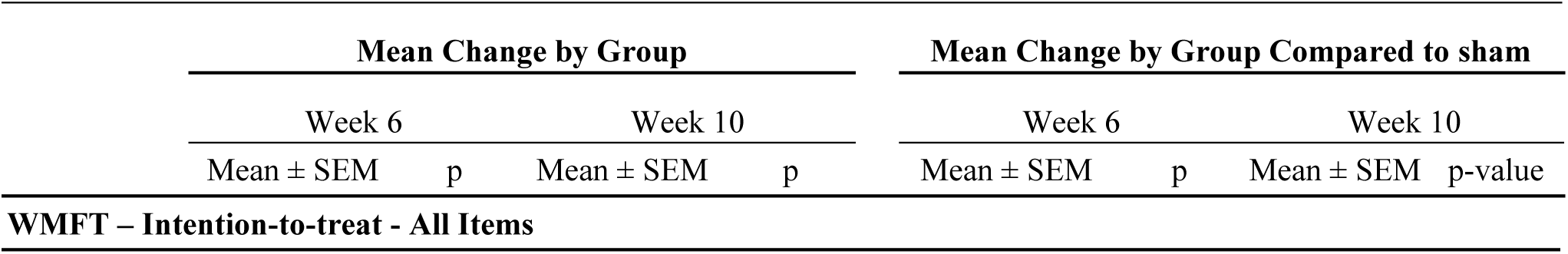
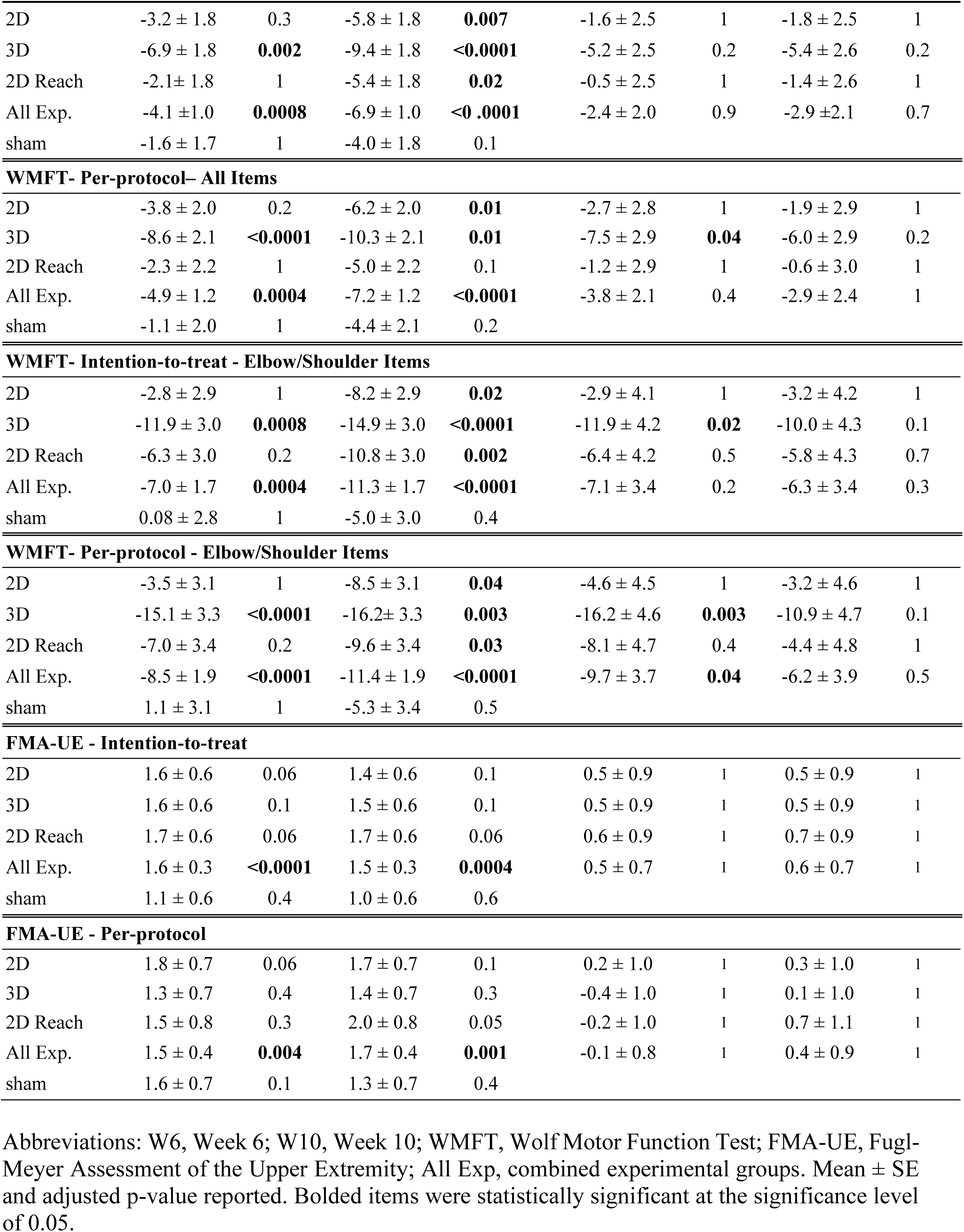
Changes in WMFT and FMA-UE at Weeks 6 and 10 from baseline, absolute change and change relative to sham (intention-to-treat and per-protocol)

**Table 3.**
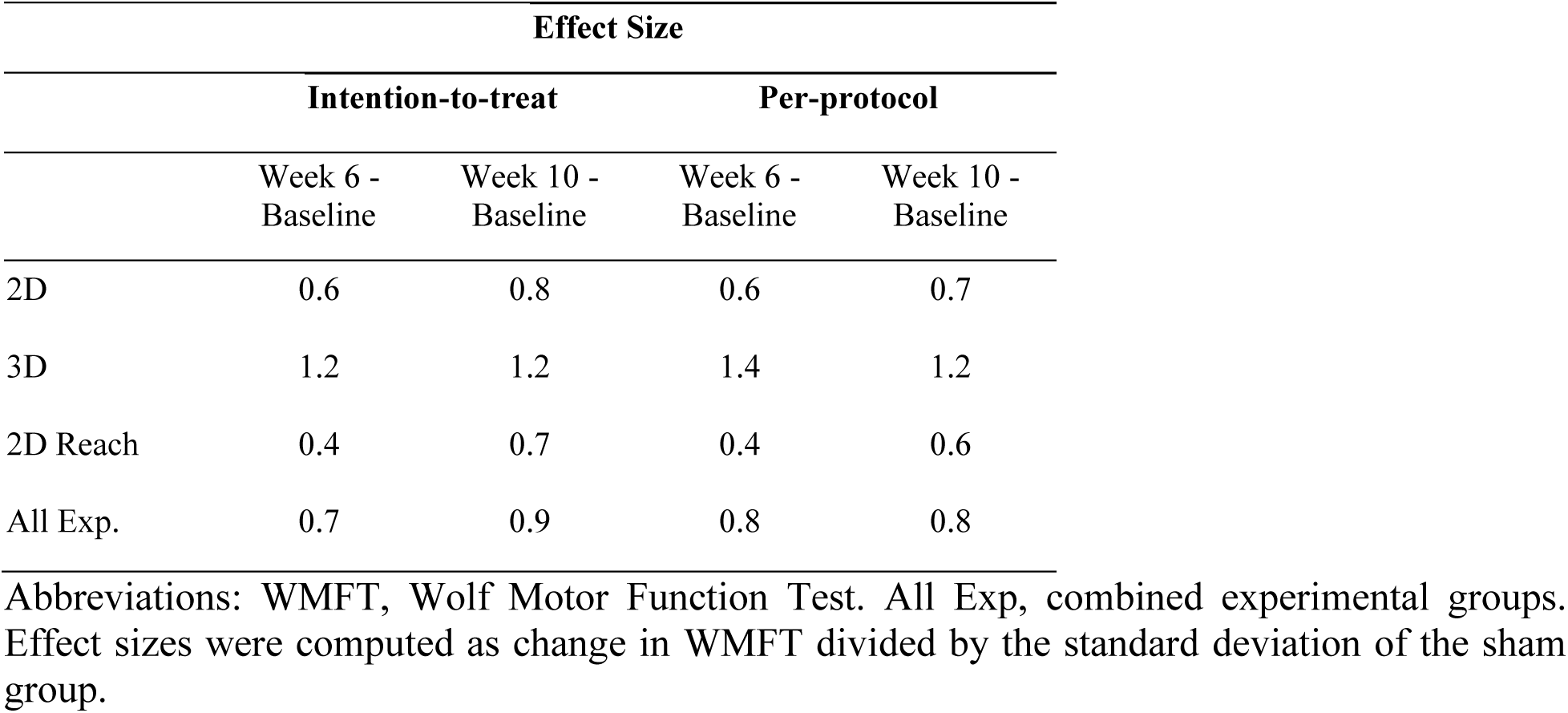
Effect size of change from baseline in WMFT at Week 6 and Week 10 for intention-to-treat and per-protocol analyses.

|  | Effect Size |  |  |  |
| --- | --- | --- | --- | --- |
|  | Intention-to-treat |  | Per-protocol |  |
|  | Week 6 - Baseline | Week 10 - Baseline | Week 6 - Baseline | Week 10 - Baseline |
| 2D | 0.6 | 0.8 | 0.6 | 0.7 |
| 3D | 1.2 | 1.2 | 1.4 | 1.2 |
| 2D Reach | 0.4 | 0.7 | 0.4 | 0.6 |
| All Exp. | 0.7 | 0.9 | 0.8 | 0.8 |

Abbreviations: WMFT, Wolf Motor Function Test. All Exp, combined experimental groups. Effect sizes were computed as change in WMFT divided by the standard deviation of the sham group.

The FMA-UE improved significantly at weeks 6 and 10 in combined experimental groups but not in the sham group in intention-to-treat and per-protocol analyses. However, this improvement was less than the MCID of 5 (Table 2). The changes in FMA-UE from baseline did not differ between any experimental group and sham. Additional secondary clinical outcomes, including MAL and MAS, did not change significantly (Table S6).

### Impact of stroke location

We also evaluated the effect of stroke location on motor recovery with MINT conditioning. People with lesions involving at least the PLIC (PLIC±) improved less than those without PLIC damage (No PLIC; p=0.02; Fig. 2D). People with lesions involving PLIC, BG, and CR had a nonsignificant trend of less improvement than those whose lesions did not involve any of those regions (however, the number of participants in the latter group was very low).

### Kinematic outcomes

For a more sensitive measure of movement changes due to MINT, we measured wrist kinematics relative to the shoulder during various reaching movements (Fig. 3A). At week 6, in the sweep task, the combined experimental groups showed a nonsignificant trend of improvement in sweep area compared to sham (difference of 201 cm²; p = 0.08, one-tailed t-test), while the 3D group improved significantly compared to sham (difference of 275 cm², p=0.03; Fig. 3A). Active range of motion (AROM) in forward reaching improved by 4 cm more than sham in combined experimental groups (p = 0.03), and the 3D group improved by 4.8 cm more than sham (p = 0.02; Fig. 3A). AROM in vertical reaching improved by 6.9 cm more than sham in combined experimental groups (p = 0.007), with the 3D group increasing by 7.4 cm more than sham (p = 0.01; Fig. 3A). In side reaching, the 3D group and combined experimental groups showed nonsignificant trends of improvement compared to sham (3.0 cm and 2.0 cm, p=0.1 and 0.2, respectively; Fig. 3)

**Figure 3.**
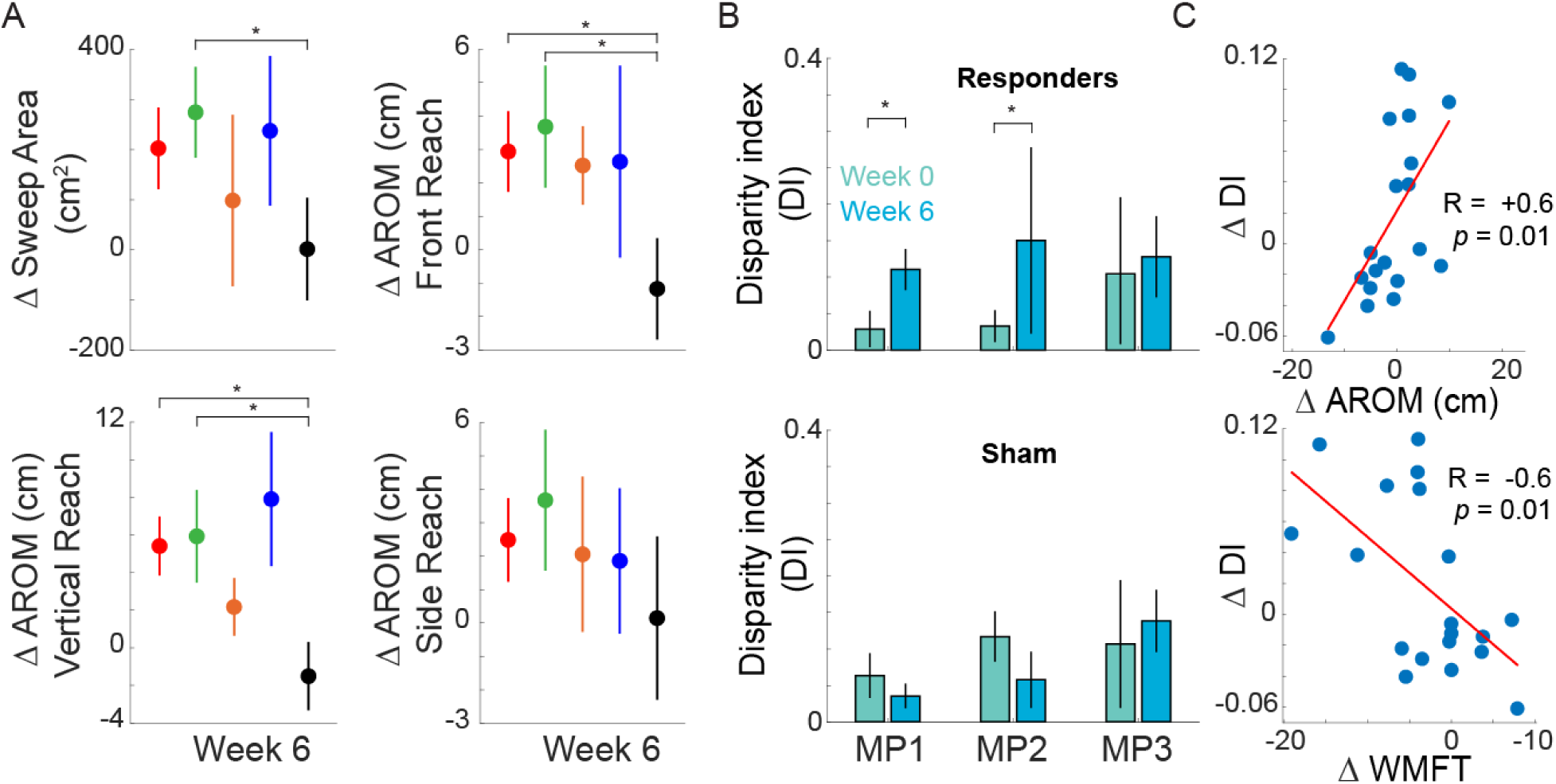
Effect of MINT conditioning on reaching kinematics and muscle synergies. (A) Mean changes on kinematic metrics at Week 6 relative to baseline for experimental and sham groups: *Top left*: Sweep area. *Top right*: Active range of motion for front reaching. *Bottom left*: Vertical reaching. *Bottom right*: Side reaching. (B) Mean (±SE) disparity index (DI) at Weeks 0 and 6 for responder (*Top*) and sham (*Bottom*) participants for each pair of trained muscles. (C) Correlations between changes in DI and changes in (*Top*) AROM during combined front and vertical reach and (*Bottom*) WMFT for combined experimental and sham groups. *, p<0.05

### Muscle synergy analysis and co-activation outcomes

After six weeks of MINT conditioning, the number and composition of synergies did not change in either responders or sham group participants. Responders showed an average of 2.6 ± 0.9 synergies at baseline and 2.4 ± 0.8 at week 6, while sham participants had 2.9 ± 0.9 and 2.8 ± 0.9, respectively. However, muscle weights within synergies did change significantly due to training. Specifically, in responders the disparity index (DI) increased significantly in muscle pairs 1 and 2 (p = 0.00004 and p = 0.005) but not pair 3 (p = 0.33; Fig. 3B). DI did not change significantly for any muscle pairs (p=0.94, p = 0.88 and p = 0.22 for muscle pair 1, 2 and 3, respectively) in sham group participants. This means that responders decreased the co-activation specifically between the muscles trained, and did not affect the other muscles in the synergy. Moreover, increased DI correlated with increased active range of motion (R = 0.6, p = 0.01) and improved WMFT (R = −0.6, p = 0.01; Fig. 3C).

## Discussion

The MINT trial is the first randomized, sham-controlled clinical trial specifically targeting abnormal muscle co-activation after stroke. The wearable MINT paradigm proved feasible, enabling at-home training that was enjoyable and motivating. MINT conditioning reduced abnormal co-activation between trained muscle sets during training. MINT conditioning also significantly enhanced arm function, while the sham intervention did not. Experimental groups combined did not improve their function significantly more than sham groups. However, this trial was designed to investigate comparisons between each MINT group and sham. Participants in the 3D group, but not the 2D groups, did improve function more than sham in the per-protocol analysis and particularly in the elbow and shoulder-related movements, which is notable because only shoulder and elbow-related muscles were trained. MINT, but not sham, conditioning also improved arm active range of motion in multiple reaching tasks. Finally, reduction in within-synergy co-activation (increased DI) during reaching correlated significantly with improvements in both function and AROM. This indicated that MINT enhanced movement by reducing abnormal co-activation rather than by overcoming long-term non-use.

The 3D group’s 6.9-s WMFT improvement, with specific shoulder and elbow items improving by 11.9 s, is clinically meaningful, approaching gains seen in constraint-induced movement therapy (10-s improvement) in less severely impaired stroke survivors (mean FMA of 43) in the subacute period. In contrast, our cohort had severe impairment (mean FMA of 16) and was enrolled at an average of 6 years post-stroke, underscoring MINT’s potential for this underserved population. Unlike vagus nerve stimulation^4^ or spinal/cerebellar stimulation, which show limited benefits in severely impaired individuals^24,25^, MINT offers a non-invasive, wearable option for those with restricted rehabilitation access. While some studies using conventional, or high-intensity^8^, occupational therapy have shown improvement in arm movement in the chronic stage of stroke^5,26^, most of those studies were performed in mildly or moderately impaired stroke survivors.

MINT effectively reduced abnormal co-activation, both during training and during reaching. This is a novel mechanism of action for stroke rehabilitation therapy, with only a few prior uncontrolled trials attempting to reduce abnormal co-activation^10,11,27,28^ or abnormal joint coupling^29^ in the past. Combined with our prior study involving in-lab myoelectric interface training, our results indicate that reducing abnormal co-activation can improve arm movement. The fact that reduced co-activation (increased DI) correlated highly with both functional gains and AROM strongly suggests that reducing co-activation led to improved movement. The fact that wrist AROM relative to the shoulder—which factors out trunk movement—did show improvement from MINT indicates that the functional improvement was not compensation, but rather true improved movement. This contrasts with many studies of task-oriented therapies in chronic stroke (though some did show evidence for reduced impairment, e.g. Queens Square, VNS-REHAB^4,7^; however, those populations were less impaired overall than our study population).

While arm function as measured by WMFT did improve significantly from MINT, secondary measures of impairment (FMA) and participant-reported function (MAL) did not. This discrepancy could be due to the fact that these are insensitive measures of improvement, especially for severely impaired individuals (i.e., nearly all of our study population). For example, significant improvement in reaching AROM could be achieved, which is functionally important to stroke survivors, without changing the FMA subscore on shoulder abduction or flexion from a 1, if the elbow is only able to be extended to 170°. The MAL is heavily biased toward items involving finger movement, which most of our participants lacked. The training in this study targeted only shoulder and elbow-related muscles and we did not expect to see improvement in more distal function, so it is not surprising that we did not see large improvements in these measures in severely impaired patients. The WMFT assessed multiple items not including the hand (i.e., shoulder and elbow items, which showed the most improvement in experimental groups vs. sham), and is a timed test, which is more sensitive to change than discrete measures.

We found that participants whose strokes involved the PLIC, a proxy for damage to the corticospinal tract, did not improve as much from MINT conditioning. This aligns with results from prior studies that analyzed effects of stroke location^22,30–32^. This may suggest that some residual corticospinal tract function is important for overcoming abnormal co-activation. Future studies will examine this question further.

This study design examined different variants of the MINT paradigm. The 3D variant’s success may stem from greater motor learning complexity or prolonged training on the most abnormally co-activating muscle sets (3 vs. 2 weeks). While the 2D Reach group did not improve as much as 3D overall, it did improve more than 2D in shoulder and elbow function. It is possible that encouraging reaching also loaded the shoulder muscles more, resulting in more co-activation^9,29^, which may have made the task more difficult. Optimizing timing of shoulder loading may be important to consider in future studies of MINT. The fact that the 2D groups did not show significantly more improvement than sham may have been due to smaller effect sizes than anticipated in these groups based on our prior study^11^. It is possible that there were more participants in this study with greater damage to the corticospinal tract than in the prior study. The greater improvement in 3D group participants suggests that longer training on the most co-activating muscle sets may have led to a greater improvement in function. Participants in this study performed about 300 repetitions per day, many more than typically done in conventional therapy (either in clinic or at home) though less total than a prior study of telerehab therapy in subacute stroke^33^. Dosage (including the number of muscle sets to train and total time to train) and intensity are important factors to consider in future studies of MINT. Although the current MINT primarily targets proximal muscles, as abnormal co-activation is most prominent in these muscle groups, it can also be applied to distal muscles, such as the wrist extensors and flexors, where abnormal synergies have been observed during reaching^34^. However, we expect that MINT will be most beneficial as an adjunct to other therapies. Combining MINT with task-based practice, as well as strategies such as behavioral contracts, homework assignments focused on daily activities, and real-life progress tracking, may enhance the integration of trained movements into activities of daily life. Furthermore, integrating MINT with neuromodulation techniques, including brain or spinal cord stimulation or pharmacologic enhancement, may enhance effectiveness by promoting neuroplasticity and facilitating motor recovery. These combined approaches hold promise for optimizing functional outcomes and promoting better transfer to daily activities.

Other limitations of the study included a relatively high withdrawal rate and non-universal enjoyment of the game. Motivation, concentration, and training intensity are critical to improving function after a stroke^35,36^. This may be a factor in improvement. While participants rated the MINT games favorably overall, some participants did have trouble using the system, as computer illiteracy was an issue for a subset of participants, as was equipment breakdown. While the withdrawal rate was relatively high (37%), the vast majority of reasons were not related to MINT conditioning or devices but rather were related to insufficient ability to adhere to appointment schedule or training and family/personal issues. Improving usability, motivation, and robustness is a goal for upcoming studies, which should help with effectiveness in real-world clinical use. The current design of the MINT device may present challenges for users. Its somewhat bulky nature and requirement to clip to electrodes could potentially hinder ease of use among stroke survivors. Addressing this concern and creating a more accessible and user-centered design will be essential for future iterations of the device.

Overall, MINT presents a promising option as a wearable, home-based treatment to enhance arm movement and function post-stroke that could improve access and complement other forms of therapy. A pilot study also suggests that MINT may similarly help leg function^27^. Benefits from reducing co-activation could perhaps increase even more if MINT were combined with other therapies to address other deficits, such as weakness and task-specific function, or to improve excitability (such as stimulation) and motor learning^37^. Future, larger trials are warranted.

## Data Availability

Data will be made available upon peer-reviewed publication.

## Acknowledgements

We thank our participants for their valuable time and effort. We thank R. James Cotton, Jeffrey Nie, and James Nie for their efforts in kinematic data recording; to Murad Alqadi and Tyler Jacobson for their assistance with recruitment and data collection; and to Torin Kovach for his contributions to game design. We also thank Michael Ellis and Veronica Rowe for training our occupational therapists on outcome evaluations.

## Source of Funding

This research was funded in part by National Institutes of Health Grants R01NS099210, R01NS112942, R01HD113270 and NSF CAREER Award (# 2145321).

## Disclosures

M.W.S. is a consultant for Iota Biosciences, Inc. The remaining authors declare no commercial relationships or conflicts of interest related to this study.

## Supplemental Material

Figures S1-S3

Tables S1–S6

## Notes

### Competing Interest Statement

MWS is a consultant for Iota Biosciences, Inc., not related to this study.

### Clinical Trial

NCT03401762

### Author Declarations

Northwestern University IRB

